# Structural and functional connectivity of the posterior default mode network is associated with sleep disturbance rather than pain in the Twin sub-study of the MAPP Research Network

**DOI:** 10.1101/2024.12.04.24317355

**Authors:** Tejaswi Sudhakar, Marija Bogic, Jonathan E. Elliott, Miranda M. Lim, Dedra Buchwald, Eric Strachan, Swati Rane Levendovszky

## Abstract

The neurophysiological underpinnings of chronic pain and sleep disturbances, which are commonly comorbid are not well understood. Investigation of common structural and functional brain network alterations of sleep disturbance in the setting of chronic pain may shed light on common mechanisms and improve specificity of treatment for these conditions. Our study examines the association between sleep parameters (including sleep duration, sleep difficulty, and sleep chronotype), pain phenotype, and brain structural and functional connectivity in individuals with chronic pelvic pain by leveraging data from the Twin Studies of the Multidisciplinary Approach to the Study of Chronic Pelvic Pain (MAPP) Research Network. Participants (n= 34 pairs) completed extensive clinical phenotyping and 3T MRI, including resting-state functional MRI (rs-fMRI), diffusion tensor imaging (DTI), and subjective sleep assessments. We observed significant differences in total sleep time (420 ± 58 min vs. 466 ± 61 min) and sleep difficulty (study-specific Likert scale 0-3: 1.35 ± 0.81 vs. 0.95 ± 0.68) among individuals with chronic pelvic pain compared to those without chronic pelvic pain, with sleep difficulty being associated with altered default mode network (DMN) connectivity. No associations were found with the salience network (SN), a network implicated in chronic pain. Additionally, increased total sleep time was associated with decreased posterior callosal white matter integrity. These findings underscore the complex interplay between chronic pain and sleep disturbances and have implications for distinct treatment approaches for patients with comorbid pain and sleep disorders.

## INTRODUCTION

Chronic pain, defined by pain lasting more than 12 weeks, often occurs alongside sleep issues, making treatment harder and increasing healthcare costs. Many people with chronic pain report sleep problems (67-88%), such as trouble falling or staying asleep and non-restorative sleep,^1^ and 50-80% of patients diagnosed with the severely disabling sleep disorders report comorbid chronic pain.^2,3^ Chronic pain conditions disrupt sleep architecture by increasing total wake time, bypassing non-rapid eye movement (REM) sleep stages, and potentially completely suppressing REM sleep.^4,5^ Sleep disruption, withal, is increasingly acknowledged as a direct contributor to pain development and has been shown as a good predictor of new-onset pain or exacerbation of chronic pain symptoms.^6,7^ In fact, lack of sleep can also make people more sensitive to pain, even resembling symptoms of fibromyalgia, showing that poor sleep can increase pain sensitivity in healthy individuals.^8,9^ Furthermore, sleep deprivation has severe implications for individuals with musculoskeletal conditions and chronic pain.^10,11^ It therefore appears that chronic pain and disrupted sleeping-waking patterns co-occur but current research does not provide conclusive evidence supporting either mechanistic synergy or independence between the two conditions. Thus, the neuropathologic underpinnings of comorbid chronic pain and sleep pathology are not well understood.

*Pathophysiology of pain*: Pain mechanisms can be divided into three main types: nociceptive, neuropathic, and nociplastic pathways.^6,12,13^ Nociceptive pain arises from tissue damage and inflammation, triggering pain through standard pain pathways. Neuropathic pain occurs due to disease or injury affecting the somatosensory nervous system, leading to nerve dysfunction.^14–16^ Nociplastic pain, a newer category, involves heightened pain and sensory responses in the central nervous system (CNS) without clear signs of tissue damage. Chronic Overlapping Pain Conditions (COPCs) is a recently coined umbrella term for non-inflammatory, peripheral-based conditions like fibromyalgia, irritable bowel syndrome, chronic fatigue syndrome, headache, temporomandibular disorder, low back pain, interstitial cystitis/bladder pain syndrome (IC/BPS), and chronic prostatitis/chronic pelvic pain syndrome (CP/CPPS).^17–19^

*Sleep pathology and its association with pain:* Impaired sleep is concretely established in the literature as a critical contributor to poor pain modulation and is mediated by loss of nociceptive inhibitory pathways, impaired cognitive processing of noxious input, and heightened inflammatory processes.^20–22^ Sleep actively facilitates functional reorganization of neuronal circuitry. In this context, disrupted sleep can significantly exacerbate an individual’s pain condition, increasing sensitivity to pain, or alternatively increasing pain interference.^4,23,24^ Pain and sleep share overlapping brain circuits, including the serotonin system, which can influence pain sensitivity and emotional responses, impacting central sensitization and the persistence of pain.^25,26^ Non-pharmacological interventions, including increasing total sleep time, cognitive-based therapy for insomnia (CBTi), relaxation, mindfulness, exercise, and physical therapies, have improved pain management and sleep-wake cycle regulation.^4,27–31^ Therefore, disruptions of shared brain networks involving pain and sleep may independently or synergistically contribute to the initiation and maintenance of chronic pain or sleep instability. Further research is needed to clarify how these two conditions interact.

*Imaging findings in chronic pain:* COPC patients and individuals with sleep complications exhibit altered cerebral functional and structural connectivity. Furthermore, central sensitization in pain also alters CNS structure and function with amplification of sensory signaling, resulting in development of somatoform disorders.^32^ A critical network implicated in pain regulation is the default mode network (DMN), a functional network of brain regions that show synchronous temporal fluctuation of their electrical and hemodynamic activity. The DMN network constitutes the medial and lateral parietal, medial prefrontal, and medial and lateral temporal cortices, bilaterally, is characterized by balanced positive and negative correlations in activity in these regions.^33^ The hippocampus, posterior cingulate cortex, and precuneus are particularly implicated in pain-related processing.^34,35^ Additionally, the salience network (SN), which includes functional activation of the insula-fronto-cingulate brain regions, is implicated in altered central nociceptive processing.^36^ Several studies have reported alterations in brain regions within the DMN, SN, and somatosensory network in chronic pain patients. Although not well-established, this reorganization is believed to hypersensitize the SN network to facilitate processing of continuous attention-demanding painful stimuli, as a compensatory response to pain.^23,37–46^ Additionally, from an anatomical perspective, structural connectivity studies in chronic pain patients have demonstrated decreased white matter integrity within the medial prefrontal cortex, nucleus accumbens, corpus callosum, bilateral cingulum, thalamocortical tracts, insula, and hippocampus, of which several brain regions are a known component of the DMN or SN.^47–52^

*Imaging findings in sleep:* Similar to findings in chronic pain, sleep-deprived individuals also show alterations in key brain networks. Functional connectivity studies in sleep deprived individuals demonstrate both decreased within network connectivity of the DMN and SN as well as decreased DMN and SN within-network connectivity.^53–55^ Additionally, several studies that have investigated white matter changes in sleep deprivation show structural alterations within the fronto-occipital, fronto- parietal, dorsolateral prefrontal cortex, longitudinal fasciculi, corticospinal tracts, and thalamic regions.^56–59^ These alterations suggest a link between disrupted DMN and SN connectivity in both chronic pain and sleep deprivation but more research is needed to understand the shared mechanisms. Few studies have explored the combined structural and functional changes in cases of both chronic pain and sleep problems, and further neuroimaging research is necessary to clarify this relationship.

*The Multidisciplinary Approach to the Study of Chronic Pelvic Pain (MAPP I) Research Network:* The MAPP Network is a multi-site study on urological chronic pelvic pain disorders i.e., Urological pelvic pain syndrome (UCPPS). It comprises of various conditions including IC/BPS and CP/CPPS that present as pelvic pain and urologic symptoms and is categorized as a mixed nociplastic COPC.^18,60,61^ UCPPS patients report increased pain with bladder distention and lower urinary tract symptoms including urinary urgency, frequency, and incontinence. The MAPP I cohort is an all-female cohort with UCPPS compared with age-matched healthy controls (HC) from the Phase I of MAPP (MAPP I).^62,63^ This dataset included subjective and objective measures of pain and subjective measures of sleep physiology. We sought to utilize this data to explore the relationship between chronic pain and sleep disturbance. This study examined urological, non-urological, and psychosocial characteristics in an all-female cohort with UCPPS compared with age-matched healthy controls (HC) from the Phase I of MAPP (MAPP I).^62,63^

We leverage the existing MAPP1 cohort, specifically, the University of Washington’s sub-study called, The MAPP Comprehensive Twin Study, to answer whether functional and structural connectivity measures within the DMN and SN network, as well as morphometric measurements, are associated with sleep measures and whether these associations are modulated by pain in a chronic pelvic pain cohort.

## METHODS

### Participant Population

Eligibility included the following: 1) UCPPS symptoms present for a majority of the time during the most recent three months qualifying a UCPPS diagnosis; 2) age ≥ 18 years; 3) a non-zero response on the bladder/prostate or pelvic pain/pressure/discomfort scale within 2 weeks; 4) consent to provide a blood sample or cheek swab to test DNA for genes related to the main study goals; and 5) absence of any condition that could cause UCPPS symptoms. Inclusion and exclusion criteria for individuals at the University of Washington are detailed in Afari et al.^63^ Additionally, for the MAPP Comprehensive Twin Study, only female twins were recruited from The Washington Twin Registries Study. The Institutional Review Board (IRB) at the University of Washington Medical Center approved this study protocol. All individuals provided informed consent. Clinical phenotyping was conducted to characterize participants with and without CPPS.

### Pain assessments

Along with general demographic and socio-economic measures, several urological measures such as Symptom and Health Care Utilization Questionnaire (SYM-Q),^64^ Interstitial cystitis symptom and problems, American Urological Association Symptom Index Score (AUASI),^65^ and the Female Genitourinary pain scale (GUPI)^66^, were collected among many others.^64^ In addition, non-urological or pain measures, including functional status (SF-12),^66^ PROMIS,^67^ The Multiple Ability Self-Report Questionnaire (MASQ),^68^ Perceived Stress Scale (PSS),^69^ and Short Form McGill Pain Questionnaire(SF-MPQ),^70^ etc. were also conducted to assign patient diagnosis as per the MAPP protocols.

### Sleep assessments

Sleep was assessed via the rMEQ,^71^ self-reported total sleep time, sleep difficulty, and Epworth Sleepiness Scale (ESS), a measure for daytime sleepiness,^72^ The study protocol includes a self-reported total sleep time and sleep difficulty on a scale of 0-3 (Question: How often do you have difficulty falling asleep or staying asleep? With 0 = never, 1 = sometimes, 2 = often, 3 = always). It also included the Epworth Sleepiness Scale (ESS) and the reduced Morning Eveningness Questionnaire (rMEQ). rMEQ determines whether the individual favors a morning or evening chronotype. Since no studies show association between pain and rMEQ, this measure was not used for any covariate analyses.

### MRI Acquisition (rs-fMRI and DTI)

For the MRI, participants were scanned on an empty bladder and then they consumed ∼350cc of water. After approximately 40 minutes, we performed a resting state fMRI (rs-fMRI) scan. The participants were rescanned with identical rs-fMRI scan parameters. During the rescan, a 3D T1 and DTI data was also acquired.

All scans were performed on a 3T Philips Achieva scanner with 32-channel SENSE reception.^73^ rs-fMRI scans were acquired with a single shot echo planar imaging (EPI) with the following imaging parameters: TR/TE = 2000/30 ms, flip angle = 77°, FOV = 220 mm × 220 mm, matrix = 64 × 64, slice thickness = 4.0 mm, slice gap = 0.5 mm, slice acquisition = ascending, 38 slices, SENSE factor = 2, number of dynamics = 300. For DTI scans, diffusion directions uniformly distributed along 64 directions at b=1000 s/mm^2^ and a reference image with b=0 s/mm^2^. Other imaging parameters were TR/TE=9500/88 ms, flip angle: 90°, FOV = 256 x 256, matrix= 128 x 128, slice thickness = 2mm, 75 slices. A 3D- T1 MPRAGE scan was acquired for registration and volumetrics with the following parameters: TR/TE = 2300/2.98 ms, TI = 900 ms, flip angle = 9°, FOV = 256 mm × 256 mm, matrix= 256 × 256, 240 slices, 1 mm thickness, SENSE factor = 2.

### Image Processing and Analysis (rs-fMRI and DTI)

All analyses were done in FSL v6.0 unless specific.^74^ The empty bladder resting state data was used for analysis. The BOLD time series was first corrected for head motion, followed by removal of low-frequency signal drifts and despiking, which was performed using AFNI (v22.1).^75–77^ The fMRI time series was skull-stripped by applying a brain mask generated from its first volume.^78^ The T1 images was corrected for bias field and B1 signal inhomogeneity corrections followed by registration to standard MNI atlas in 2mm space. The first volume of the BOLD times series was extracted and registered to the native T1 space.^79–81^ Next, the transformation matrices for converting from BOLD space to T1 space and T1 space to MNI space^74,80,81^ were combined to obtain a single transformation of the BOLD series into MNI space. Finally, the data was smoothed using a full-width half max (FWHM) of 6 mm. The data was then processed to obtain group-level resting state networks and identify default mode network (DMN) and salience network (SN). These two networks were chosen since they are both implicated in chronic pain. Once identified, we performed dual regression^82,83^ to obtain DMN and SN maps for each individual. We then used generalized linear model analysis to determine whether resting state network connectivity in these networks differed between individuals with and without pain and whether they were associated with sleep measures. The inter- correlatedness due to the twin participants was adjusted for in the model. All results were determined using threshold-free clustering enhancement followed by a correction for multiple comparisons using false discovery rate of p=0.05.

For the DTI data, the b=0 image was separated from the remaining 64 DWI images. DTI data was corrected for motion and eddy current distortions in FSL and skull-stripped.^78,84–86^ Following skull-stripping, fractional anisotropy (FA) and mean diffusivity (MD) values at the voxel level were calculated in FSL. We used tract-based spatial statistics (TBSS) to determine whether white matter tracts differed between individuals with and without pain and whether their integrity was associated with sleep measures.^87^ All results were determined using threshold-free clustering enhancement^88^ followed by a correction for multiple comparisons using false discovery rate of p=0.05.

## RESULTS

### Patient Demographics

The Twin Study component of MAPP 1 recruited and scanned 34 twins (37 ±16 years old, all female). Of those, 30 pairs completed their rsfMRI, and only one twin of the remaining 4 completed the rsfMRI. Twenty-one pairs (37 ±16 years old, all female) completed their DTI, and only one of the 8 pairs completed the DTI. There were fewer completed DTI scans because they were collected post-bladder-filling, and not everyone who completed the pre-bladder-filling scan completed the post- filling scan. Of the 30 twin pairs, where both individuals completed the study, 12 pairs were concordant for the absence of pain, 2 pairs were concordant for pain, and 16 pairs were discordant for pain. Of the remaining 4 twin pairs, where only 1 of the twins completed the study, 3 were diagnosed with pain.

### Sleep Measures

Considering the pain phenotype in all individuals (including twins), total sleep time in individuals without pain was 466 ± 61 minutes compared to 420 ± 58 minutes in individuals with pain (p=0.006; **Figure 1A)**. Similarly, sleep difficulty scores in individuals without pain were 0.95 ± 0.68 and 1.35 ± 0.81 in individuals with pain (p=0.04; **Figure 1B**). Epworth Sleepiness Scale (**Figure 1C)**. When the participants were divided into twin pairs where both individuals had pain, both individuals had no pain, or one individual had pain, and one did not, no significant differences were observed in any sleep-related measurement.

**Figure 1:**
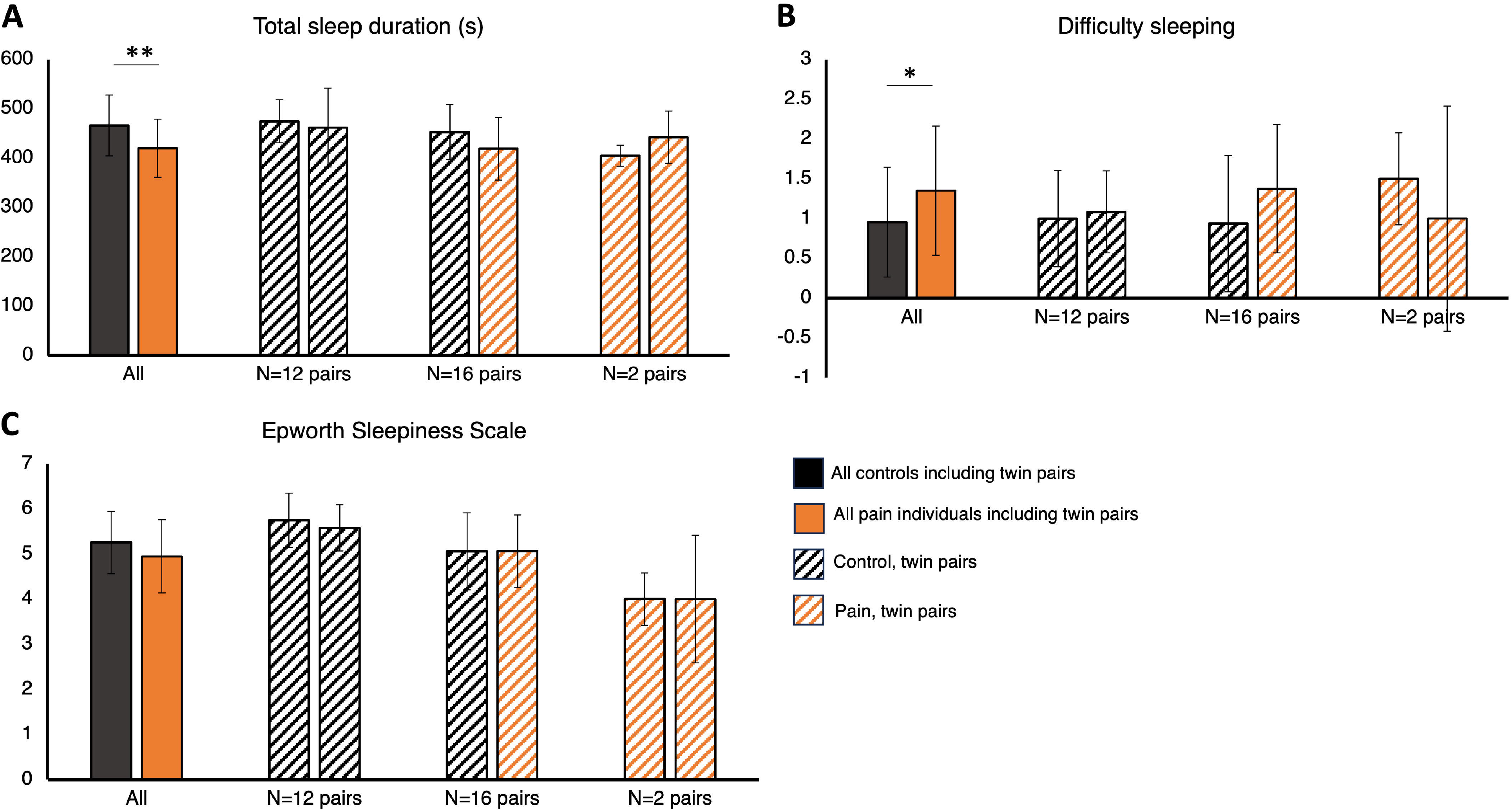
Total sleep duration, MEQ, difficulty sleeping, and Epworth Sleepiness Scale comparison between all individuals and twin pairs. Significant differences were found in total sleep duration and self-reported sleep difficulty between pain individuals and healthy controls (Figure 1A**,B**). No other group comparisons showed significant differences in total sleep duration. No significant differences were found in Epworth Sleepiness Scale scores between any groups (**1C**).

### Resting State Functional Magnetic Resonance Imaging

All resting state networks were identified and depicted in **Supplementary** Figure 1. **Figure 2** shows the DMN and SN networks used for analyses. No differences were detected between the group with pain and the group without pain. Only sleep difficulty was associated with DMN connectivity (p<0.03). A higher score on sleep difficulty (greater difficulty) was associated with lower connectivity in DMN. No significant observations were detected for SN.

**Figure 2:**
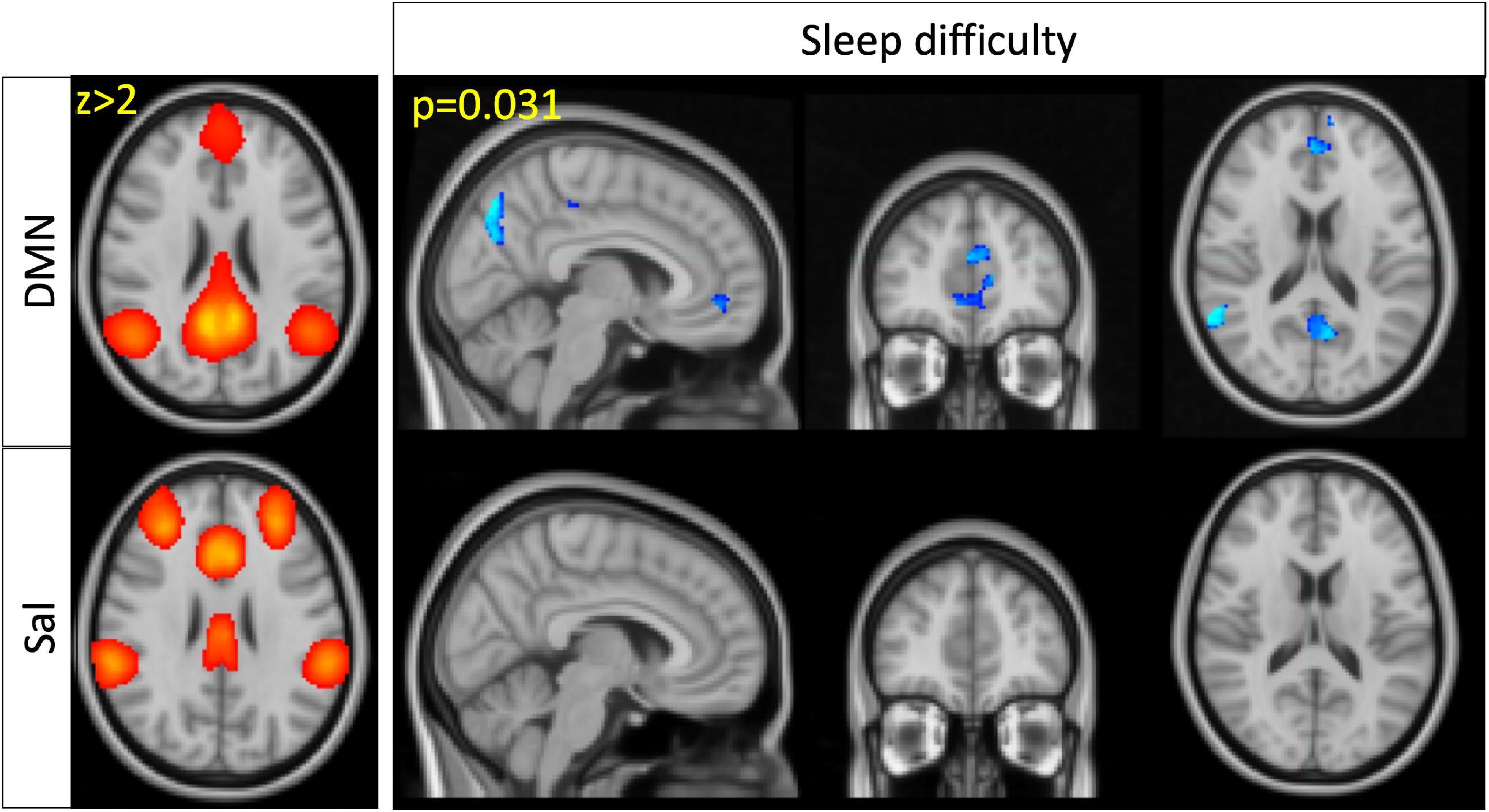
Functional connectivity assessment between pain individuals and healthy controls using the DMN and Salience network as seed points used for analysis. No differences were detected between pain individuals and healthy controls. Increased sleep difficulty was found to be associated with decreased DMN connectivity. No differences were found associated within the salience network.

### Diffusion Tensor Imaging

**Figure 3** shows the FA maps for representative individuals (top row) and at a group level (middle row). With TBSS, no differences were detected between the group with pain and the group without pain. The mean skeletonized FA is overlaid in green on top of the group-averaged FA map. Only total sleep time was associated with the white matter integrity (bottom row). This observation was in the mid-cingulate and retrosplenial cingulum i.e., a shorter total sleep time was associated with decreased integrity in these regions.

**Figure 3:**
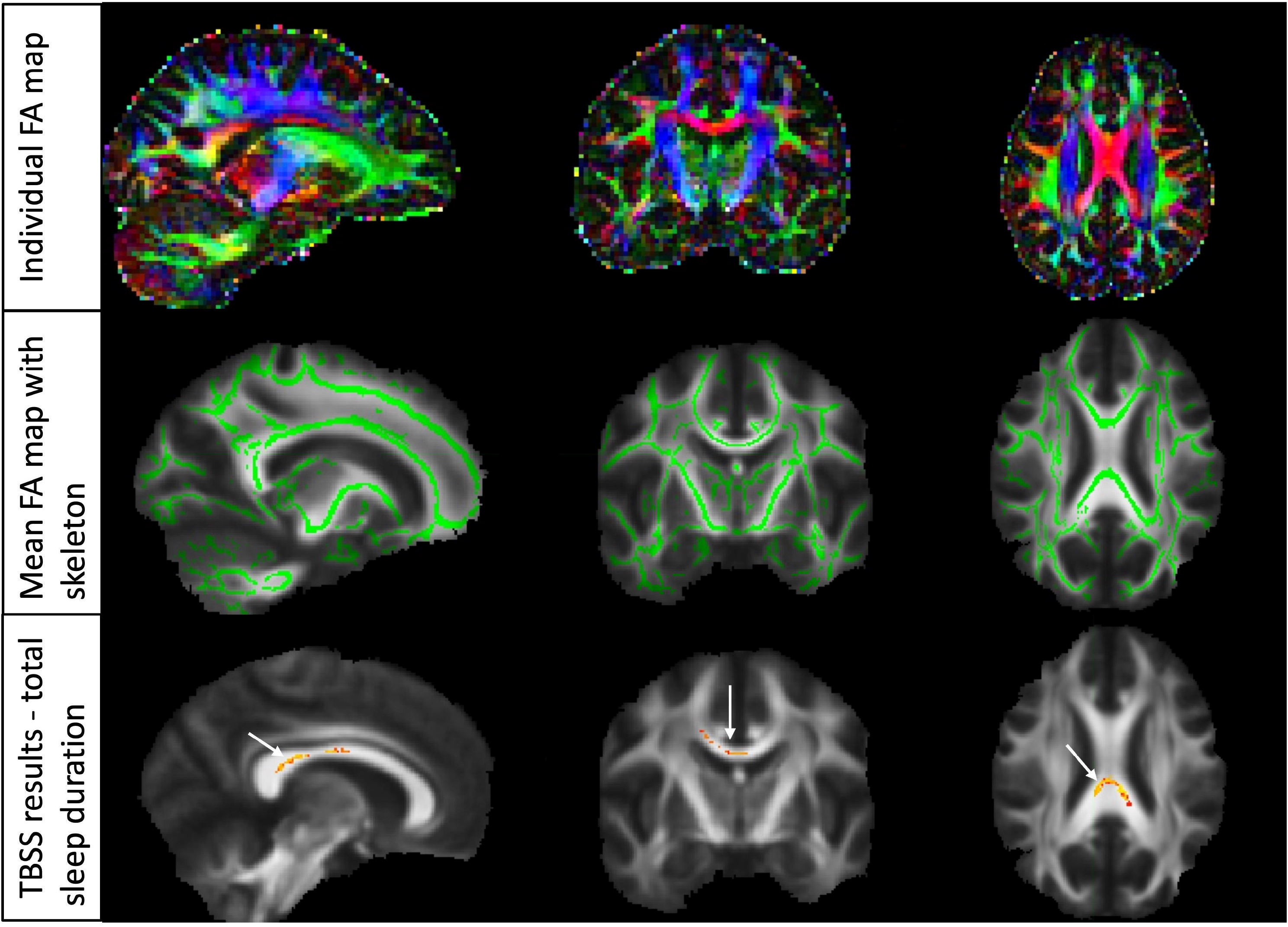
Structural connectivity assessment between pain individuals and healthy controls using diffusion tensor imaging (DTI). A representative fractional anisotropy (FA) map of an individual is shown in the first row. A representative fractional anisotropy (FA) map at the group level is shown in the second row. The third row shows TBSS analysis demonstrating no differences between pain individuals and healthy controls. Decreased sleep duration was associated with decreased white matter integrity within the mid-cingulate and retrosplenial cingulum.

## DISCUSSION

This study examined how sleep quality and duration, based on self-reported measures, relate to chronic pelvic pain and brain connectivity patterns. Our findings were (1) People with chronic pelvic pain reported shorter sleep duration and more sleep difficulties compared to those without pain. (2) Difficulty sleeping was linked to changes in brain network connections, specifically in the default mode network (DMN). The usual negative correlation between the DMN and the salience network wasn’t observed here, suggesting an atypical pattern in those with sleep issues and pain. (3) Reduced sleep duration was associated with lower white matter integrity, particularly in brain areas involved in processing emotions and self-reflection, like the cingulum. However, there was no connection between white matter integrity and pain severity.

This study confirms that people with chronic pelvic pain tend to sleep less and experience more sleep difficulties—findings that align with previous research. For example, a large study from Vienna found that over 30% of individuals with chronic pain had trouble falling asleep, and more than 60% experienced fragmented sleep.^89^ It is well demonstrated in the literature that UCPSS has a high incidence of concomitant presentation of non-urologic symptoms, such as sleep difficulty, attributed to its underlying centrally sensitization central sensitization, where pain pathways in the brain become overly responsive.^27,37,90–92^ This sensitization can disrupt sleep and keep the body in a prolonged state of hyperarousal, intensifying the stress response in those with chronic pain.^15,93–95^ Our observation of decreased total sleep duration and increased sleep difficulty in individuals with chronic pelvic pain supports this hypothesis indicating that hyperarousal and pain perception synergistically contribute to chronic pain processing networks. One drawback of our study is that the sleep measures are subjective and introduce heterogeneity. Future studies that utilize objective measures of sleep-wake patterns can allow better delineation of the various features of sleep architecture.

We did not observe a significant difference in ESS scores. However, other pain studies report greater daytime sleepiness in chronic pain participants. A study in chronic neck pain showed that day time sleepiness was greater in individuals with pain than those without.^96^ Similar observations are also made in musculoskeletal pain.^97^ Other studies in chronic back pain report a comparable lack of discrimination in daytime sleepiness between individuals with chronic back pain and controls.^98^ However, not many individuals experience daytime sleepiness. Poor sleep quality at night may manifest as daytime sleepiness that the ESS captures. This scale, however, is subjective and sleepiness during the day can relate to other physical and mental health conditions. Furthermore, analgesic medications including narcotics may increase daytime sleepiness with subsequent adverse effects experienced during nighttime sleep.^92,99^ As such, there may only be a loose association with chronic pain.

Our second finding, from resting-state functional connectivity analysis, showed that in patients with chronic pelvic pain, increased sleep difficulty was linked to lower connectivity in the default mode network (DMN), particularly in the medial prefrontal cortex, parietal, precuneus, and posterior cingulate regions. However, this DMN connectivity was not influenced by pelvic pain alone. Additionally, we found no significant connection between sleep measures (duration or difficulty) and the salience network (SN). Similar findings are also reported in individuals with poor sleep or after sleep deprivation.^53,54,59,100–102^ For instance, Kilgore et al. found a negative relationship between DMN connectivity and total sleep time in people with insomnia,^103^ and De Havas et al. reported decreased DMN activity after sleep deprivation.^104^ While poor sleep and pain are related, and hypersensitivity in chronic pain often involves the anterior cingulate, insula, and prefrontal cortex (key regions of the SN), we did not observe the typical decrease in DMN connectivity related to pain or the usual inverse relationship between the DMN and SN.^41,44,105–109^ We discuss possible reasons for this lack of association with pain and SN connectivity in the paragraphs to follow.

Our third finding showed a link between sleep duration and the structural health of white matter, especially in certain fibers of the corpus callosum (posterior body and splenium) that connect the two brain hemispheres. These areas play a role in cognitive and emotional processing, self-awareness, pain regulation, and sleep control.^110,111^ The splenium likely connects the lateralized DMN regions which showed reduced connectivity with poor sleep, confirming a structure-function relationship. Although previous studies suggest a connection between corpus callosum integrity and pain, we did not find this link with pain in our study possibly due to similar reason as the rs-fMRI.^112–115^ Poor sleep quality is related to a longitudinal atrophy of the corpus callosum. Several studies have found decreased integrity of the splenium in insomnia and sleep-deprived individuals.^102,116^ Furthermore, the callosal integrity is critical for the coordination of slow-wave sleep.^117^ There is an inverted U-shaped association between callosal integrity and sleep duration, where both shorter as well as longer sleep duration is associated with atrophy of the callosum.^118,119^

Finally, several twin studies have demonstrated a relationship between pain and sleep disturbance. In a study of more than 2000 twin pairs, authors demonstrated that sleep quality and low back pain were moderated not only by the same genetic and environmental factors but were even more pronounced in monozygotic twin pairs.^120^ Another study of 400 twins reported that sleep quality, pain, and depression had heritability estimates of 37%, 25%, and 39%, respectively.^121^ A study of more than 1200 participants that assessed rs-fMRI and genotype data in chronic pain and sleep disturbance demonstrate functional connectivity changes within the prefrontal, temporal, precentral and post-central, anterior cingulate, and fusiform cortices, as well as within the hippocampus.^37^ Although our study cohort included pain- concordant and pain-discordant twin pairs and healthy controls, the number of twin pairs are insufficient for analyses to assess effects of genetics or environment or both. Our analysis was designed to assess sleep architectural differences between UCPPS and healthy controls. In this regard, we treated twinness as a nuisance variable, likely reducing the power of the analysis. Therefore, our lack of results between pain and no-pain or lack of association with DMN and SN may be due to the small sample size and multiple comparisons corrections for twinness that rendered the pain-related differences in DMN between twins unobservable.

## CONCLUSION

In conclusion, our study highlights the relationship between sleep parameters, pain phenotype, and brain connectivity measures in individuals with chronic pelvic pain. We observed significant differences in total sleep duration and sleep difficulty among those with pain, with sleep difficulty being associated with altered DMN connectivity. No associations were detected with pain which could be due to the twinness in our data. Our results emphasize the importance of considering sleep quality and brain connectivity in the assessment and management of chronic pain conditions like pelvic pain.

## Data Availability

All data produced in the present study are available upon reasonable request to the authors.

