## Supplementary figure for "Structural and functional connectivity of the posterior default mode network is associated with sleep disturbance rather than pain in the Twin sub-study of the MAPP Research Network"

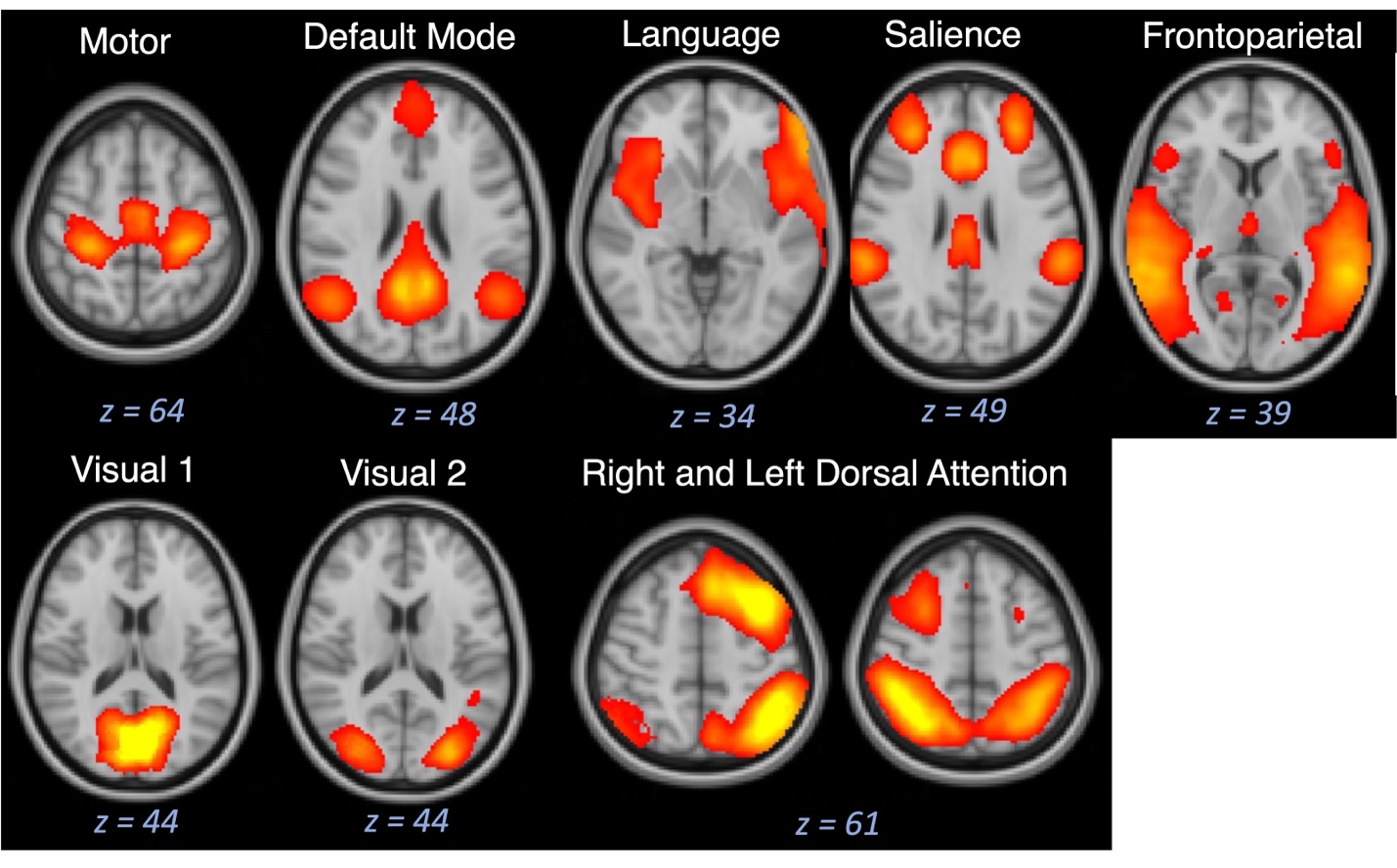

**Supplementary Figure 1:** All resting state functional connectivity networks identified. We identified the following resting state networks within our chronic pelvic pain cohort: motor network, default mode network, language network, salience network, frontoparietal network, visual network 1, visual network 2, right dorsal attention network, and left dorsal attention network.
